# Alpha thalassemia/mental retardation X-linked (ATRX) protein expression in human pituitary neuroendocrine tumours and its reported correlation to prognosis and clinical outcomes: A systematic review

**DOI:** 10.1101/2024.10.25.24316113

**Authors:** Edward Wang, Fabio Rotondo, Michael D. Cusimano

## Abstract

Mutations in Alpha thalassemia/mental retardation X-linked (ATRX) have been implicated in several cancers, including gliomas, sarcomas, neuroendocrine tumors, and other mesenchymal malignancies. ATRX loss contributes to oncogenesis, accelerates tumor growth, and reduces survival by disrupting epigenetic and telomere mechanisms. Additionally, ATRX loss can increase tumor sensitivity to treatment therapies. While research has explored ATRX expression in many cancers, data on its relationship to prognosis in pituitary neuroendocrine tumors (PitNETs) remain inconsistent. This systematic review aims to summarize all available studies on ATRX mutations and expression in PitNETs. A systematic search of PubMed, Scopus, and EMBASE databases was conducted to identify publications between 2014 and 2025 that investigated ATRX mutations or expression in PitNETs, following PRISMA 2020 guidelines. Of 32 identified studies, ten met the inclusion criteria, covering a total of 513 PitNETs. Only 20 tumors (3.9%) showed a loss of ATRX expression. Among these, 60% exhibited corticotropic pathology, while 20% displayed lactotrophic pathology. A small subset of tumors (30%) was classified as pituitary carcinomas with aggressive and proliferative characteristics. Additionally, 10% demonstrated the alternative lengthening of telomeres (ALT) phenotype, 50% had concurrent TP53 mutations, and 25% had elevated Ki-67 indices, indicating a higher proliferative index. Although ATRX mutations are rare in PitNETs, tumors with ATRX loss tend to be more aggressive and exhibit proliferative and transformative properties. Due to the limited number of cases, further studies with larger, prospective cohorts are needed to better understand the role of ATRX loss in PitNET progression and aggressiveness.

## Introduction

Alpha thalassemia/mental retardation X-linked (ATRX) is an ATP-dependent chromatin remodelling protein within the switch/sucrose nonfermentable (SWI/SNF) family of chromatin remodelling proteins. ATRX features two conserved domains: an ATRX-DNMT3-DNMT3L (ADD) domain at the N-terminus and a SNF2 ATPase/helicase domain at the C-terminus. The ADD domain contains critical structures for chromatin localization and binding, housing a GATA-like domain and a PHD-like domain [1]. In neuronal cells, a MECP2 binding domain found within the ATPase domain drives recruitment of ATRX to heterochromatin targets. The ATP-dependent helicase domain is characteristic of other SNF chromatin remodeling proteins and enables functional activity of ATRX (Fig 1) [2]. Germline mutations in these domains are associated with the development of ATR-X syndrome, from which the protein derives its name [1].

**Fig 1.**
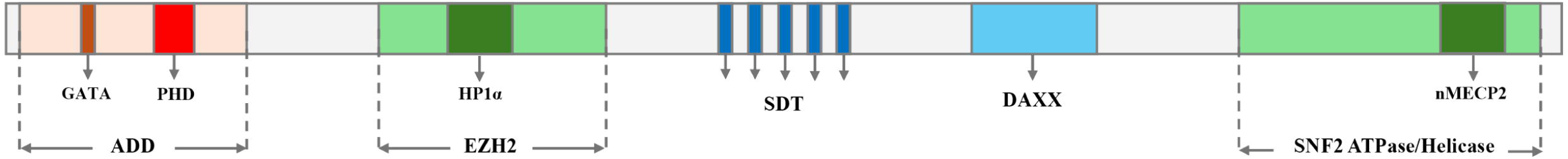
Schematic representation of the ATRX protein showing its major functional domains. The N-terminal ADD domain, containing GATA-like and PHD-like domains, mediates chromatin localization and binding [1,2]. A neuronal MECP2 (nMECP2) binding domain provides an alternative route for ATRX recruitment to chromatin targets [2]. The unstructured central region harbors “SDT-like” domains, which modulate MRN activity, and a DAXX-binding domain essential for DAXX-mediated heterochromatin regulation [3]. Towards the C-terminus, the SNF2 ATPase/helicase domain promotes chromatin remodeling. Additional features include an EZH2 domain, which supports PRC2-associated histone methylation, and an HP1α binding domain, facilitating heterochromatin recognition and PML nuclear body interactions [1,2].

ATRX also contains three other domains critical to its function: (1) an enhancer of zeste homolog 2 (EZH2) binding domain, which includes an HP1α binding domain, (2) several central “SDT-like” domains within the unstructured central region of the protein, and (3) a death domain-associated protein (DAXX) binding domain. The ATRX/EZH2 complex is recruited to a larger polycomb repressive complex 2 (PRC2) to modulate trimethylation of H3K27 (H3K27me3), silencing genes surrounding the modified histone (Fig 2). This function is utilized in transcription regulation as early as embryonic development, where ATRX is involved in X-chromosome inactivation [1,2]. The ATRX/DAXX complex localizes to promyelocytic leukemia nuclear bodies (PML-NBs), subnuclear multiprotein structures involved in gene expression, chromatin regulation, and DNA repair [2,4,5]. ATRX/DAXX modulates the deposition of histone H3.3 at guanine-rich heterochromatin regions with highly repetitive elements, including telomeres and pericentromeric regions [1,5–7]. HP1α facilitates ATRX/DAXX preferential recognition and binding to H3K9me3, facilitating histone H3.3 deposition and maintenance of heterochromatin integrity [5,8,9]. The SDT-like domains consist of spaced repeats of Ser-X-Thr motifs, where X is either Asp or Glu. When phosphorylated, these SDT-like domains enable ATRX to interact with the MRE11-RAD50-NBS1 (MRN) complex, a necessary interaction for ATRX-mediated DNA damage response (DDR) for double-stranded breaks (DSBs) and stalled replication forks [2,3]. ATRX sequesters MRN from telomeres, preventing aberrant DDR activation and preserving telomere stability (Fig 2).

**Fig 2.**
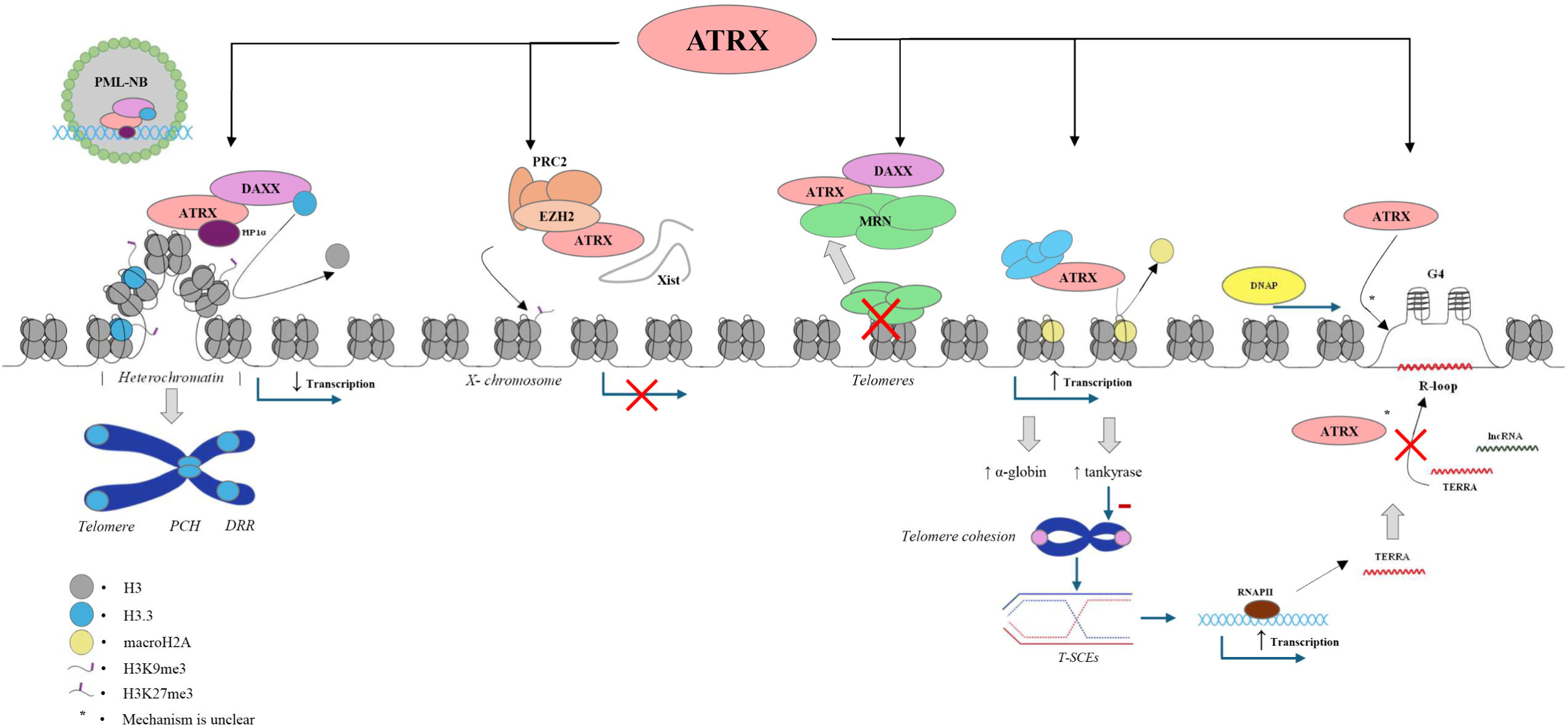
Overview of known molecular pathways of ATRX. The ATRX/DAXX complex localizes to PML nuclear bodies, where it is recruited to telomeres, pericentromeric heterochromatin, and other DNA repeat regions. This recruitment is facilitated by HP1α, which recognizes H3K9me3 marks, allowing ATRX/DAXX to deposit histone H3.3 and maintain heterochromatin integrity [1,2,5,8,9]. ATRX also forms a complex with EZH2 and is incorporated into the larger PRC2 complex, interacting with Xist lncRNA to promote X chromosome inactivation during early female embryonic development [1,2]. In addition, ATRX sequesters the MRN complex from telomeric regions, preserving telomere stability [2]. Beyond its role in heterochromatin maintenance, ATRX modulates replacement of histone macroH2A from gene loci, thereby maintaining a euchromatic state at target genes, including those encoding α-globin and tankyrase, and thus supporting their transcription. ATRX is also implicated in the DNA damage response at stalled replication forks through multiple mechanisms. It indirectly reduces telomere cohesion and telomere sister chromatid exchanges (T-SCEs) by regulating tankyrase, which in turn decreases transcription of TERRA, a long noncoding RNA (lncRNA) originating from telomeric DNA. Excess TERRA promotes secondary structures such as G-quadruplexes (G4) and R-loops that can stall replication forks [2,10–15]. ATRX helps sequester TERRA and participates in resolving these secondary structures, although its precise mechanism of action remains to be fully elucidated. Influenced and adapted from [1,2]

Beyond its role in DNA repair and gene silencing, ATRX also facilitates transcription of key genes, including α-globin, a protein essential for hemoglobin function, and tankyrase, a group of enzymes belonging to the poly(ADP-ribose) polymerase (PARP) superfamily. Impaired α-globin transcription is associated with presentation of α-thalassemia, and in germline situations, ATR-X (Fig 3) [10–12]. Tankyrase inhibits telomere cohesion and recombination between sister telomeres, which if allowed to progress, destabilize telomeres and promote aberrant transcription. The resulting transcripts form the long noncoding RNA (lncRNA) TERRA, which can hybridize to other genomic regions and induce the formation of G-quadruplexes (G4) and R-loops (Fig 2). Impaired transcription of tankyrase may prevent proper resolution of these secondary structures, leading to stalled replication forks error-prone DDR pathways [2,13–15].

**Fig 3.**
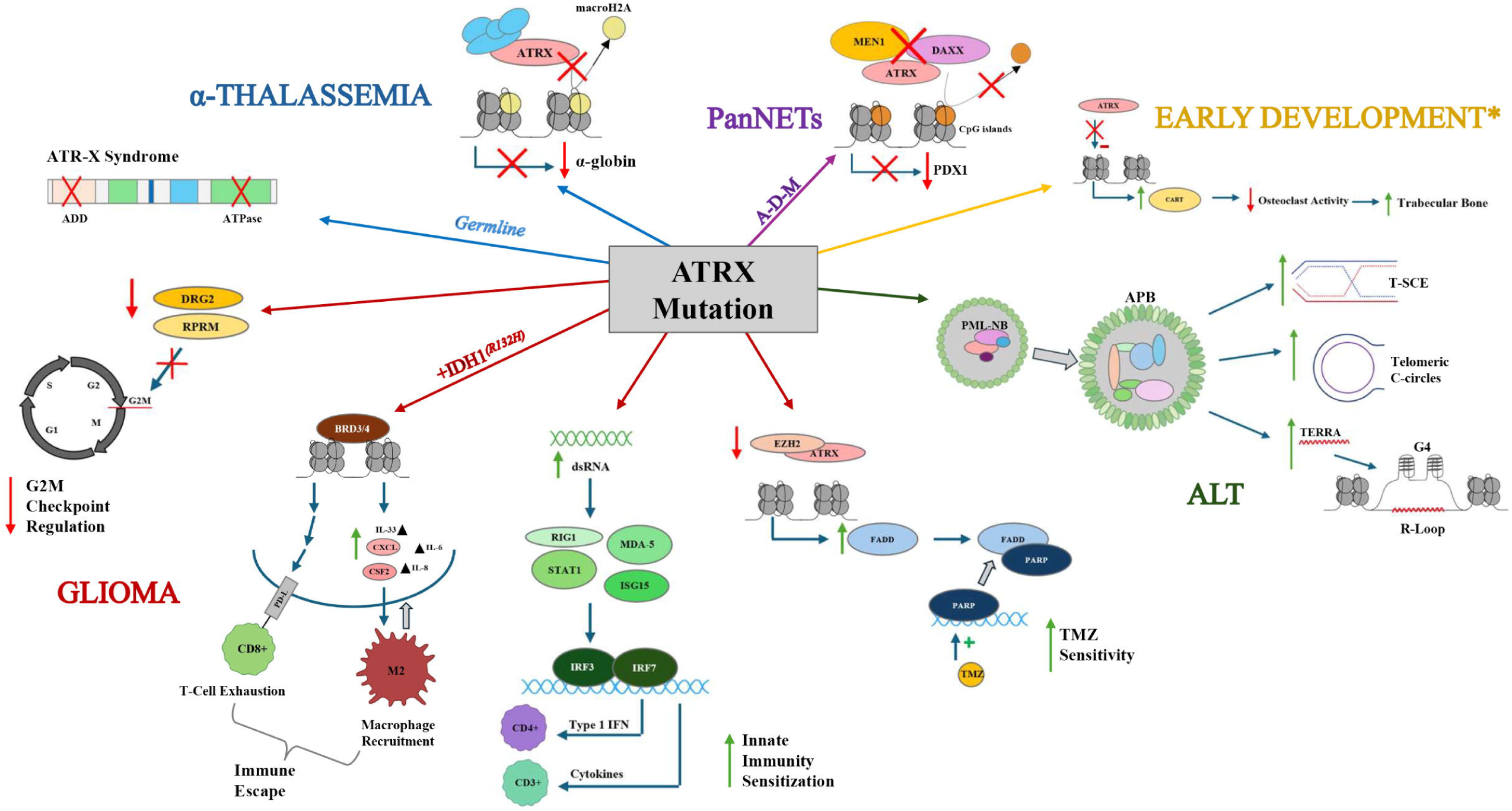
Summary of the effects of ATRX mutations on disease, tumors, and development. Mutations in conserved ATRX domains impair α-globin transcription, leading to α-thalassemia. Germline mutations of these domains induce the development of ATR-X syndrome. Mutations to ATRX in gliomas have been observed to inhibit production of DRG2 and RPRM, proteins critical for regulation of the G2M checkpoint [2,16,17]. IDH1^R132H^+ gliomas with ATRX mutations exhibit BRD3/4-dependent immune escape, which involves elevated PD-L expression and increased production of cytokines. including but not limited to CXCL, CSF2, IL-6, IL-8, and IL-33. This culminates with increased recruitment of M2 macrophages and T-cell exhaustion, facilitating immune escape [1,18,19]. ATRX loss increases dsRNA production, activating a signaling cascade through RIG1, MDA-5, STAT1, and ISG15 that boosts type I interferon and cytokine output, ultimately enhancing recruitment of CD3+ and CD4+ T cells and increasing innate immunity sensitization [1,18]. Disruption of ATRX/EZH2 complex formation reduces suppression of FADD production, allowing FADD-mediated PARP sequestration and heightening sensitivity to temozolomide (TMZ) [2,20,21]. In PanNETs, ATRX-DAXX-MEN1 (A-D-M) mutations hypermethylate CpG islands at PDX1 gene loci, compromising β-cell function [1,2,22]. Loss of ATRX is also a common driver of the ALT (alternative lengthening of telomeres) phenotype, which features APB (ALT-associated PML bodies) formation, increased telomeric sister chromatid exchanges (T-SCEs), accumulation of extrachromosomal C-circles, and heightened TERRA-induced G-quadruplex and R-loop formation [1,2,13–15]. Finally, ATRX dysfunction has been proposed to influence early bone development by downregulating CART, thereby reducing osteoclast activity and increasing trabecular bone formation [23]. Influenced and adapted from [1,2].

ATRX also participates in the resolution of G4 and R-loop structures. Although the exact mechanism of action is unclear, ATRX has been proposed to act directly or exert secondary effects that facilitate homologous recombination (HR)-mediated resolution of secondary structures. ATRX has also been proposed to inhibit the formation of secondary structures by sequestering free TERRA [1,2,24,25]. When ATRX function is disrupted, its silencing activity is lost, resulting in elevated EZH2 and MRN activity at target sites and reduced suppression of ATRX/DAXX-regulated regions. This deregulation promotes aberrant transcription [8].

Examination of the genetic factors influencing tumour initiation and progression has long been a central focus of oncology research (Table 1). Deleterious mutations disrupting ATRX function are frequently observed among several tumour types (Fig 3). ATRX mutations have been most frequently linked to the development of the alternative lengthening of telomeres (ALT) pathway, a telomerase-independent mechanism that confers replicative immortality on cancer cells [19]. Mutations affecting the DAXX and SDT-like domains promote the formation of ALT-associated PML-NBs (APBs), characteristic of ALT-positive cells. Unlike PML-NBs, APBs localize at normally silent repetitive regions, driving aberrant transcription of telomeric and pericentric sequences normally silenced by ATRX. Consequently, telomere sister chromatid exchanges (T-SCEs), extrachromosomal telomeric C-circles, and TERRA-driven G4 and R-loop structures become more prevalent, increasing the frequency of stalled replication forks. Since ATRX also participates in replication fork repair, ALT-positive cells exhibit higher rates of replication fork collapse and greater reliance on non-homologous end-joining (NHEJ), elevating mutation rates and genomic instability [1–3].

**Table 1.** Reviews of ATRX in Other Tumours.

| Author | Year | Tumour Type | ATRX Status Examined | Effect of ATRX on Outcome |
| --- | --- | --- | --- | --- |
| Gonzales-Céspedes (Navarro)[37] | 2025 | Neuroblastoma | Loss | Poor outcomes: very low survival |
| Boehm (Tothill)[38] | 2024 | Phaeochromocytoma and paraganglioma | Loss | Poor outcomes: ALT, G2M cell cycle control, lncRNA dysregulation<br>Good outcomes: increased sensitivity to treatment |
| Levine (Hawkins)[39] | 2024 | Pediatric low-grade glioma | Loss | Poor outcomes: aggressive behaviour |
| O'Neill (Rodriguez-Galindo)[40] | 2024 | Pediatric adrenocortical carcinoma | Loss | Poor outcomes |
| Sonnen (Brosen)[30] | 2024 | PanNENs | Loss | Poor outcomes: telomere dysfunction, recurrence rate |
| Chang (Demirci)[26] | 2023 | Conjunctival Melanoma | Loss | Poor outcomes: ALT |
| Hackeng (Dreijerink)[41] | 2023 | Insulinoma | Loss | Poor outcomes: aggressive behaviour |
| Haddox<br>(Riedel)[42] | 2023 | Bone and soft tissue<br>sarcoma | Loss | Good outcomes: increased sensitivity to<br>inhibitors |
| Luchini<br>(Scarpa)[31,43] | 2023 | PanNENs | Loss | Poor outcomes: ALT, metastasis, tumour progression |
| Metu<br>(Rahman) [43] | 2023 | Astrocytoma | Loss | Poor outcomes: ALT<br>Good outcomes: improved progression-free survival |
| Hackeng<br>(Heaphy)[29] | 2022 | PanNETs | Loss | Poor outcomes: ALT, decreased recurrence-free survival |
| Ren (Li)[28] | 2018 | Sarcoma | Loss | Poor outcomes: ALT, progression-free survival |
| Karsy<br>(Colman)[44] | 2017 | Glioma | Loss | Poor outcomes: ALT, astrocytoma/oligodendroglioma classification |

ALT = alternative lengthening of telomeres, PanNENs = pancreatic neuroendocrine neoplasms,
PanNETs = pancreatic neuroendocrine tumours

Loss of ATRX function correlates with increased ALT activation in melanomas and sarcomas, conferring heightened proliferative capacity and poorer disease-free survival [1,26–28]. Somatic ATRX mutations are also associated with reduced disease-free survival, elevated genomic instability, and increased tumor mutation burden in gliomas and pancreatic neuroendocrine tumors (PanNETs)[29–31]. In gliomas, ATRX dysfunction compromises cell-cycle checkpoints, fosters immune evasion, and worsens survival outcomes (Fig 3) [16,32]. Notably, metastatic progression of pheochromocytomas and paragangliomas (PPGLs) have been associated with ATRX mutations, which are among the most frequent gene-level events driving PPGL metastatic potential [17,33,34]. Reflecting these findings, the fifth edition of the WHO classification of Tumours of the Central Nervous System (2021) includes ATRX status as part of the recommended diagnostic workup for gliomas. Recent studies in PanNETs have shown that ATRX inactivation correlates with shorter disease-free intervals, more frequent ALT activation, and increased mortality [30,31,35,36]. Mutations in ATRX, DAXX, and MEN1 (A-D-M mutants) are particularly detrimental, leading to poorer prognoses and lower disease-free survival, with or without ALT phenotype progression. One proposed mechanism involves hypermethylation of CpG islands and subsequent repression of *PDX1*, driving beta pancreatic cells toward an alpha lineage, which is correlated to worse clinical outcomes (Fig 3) [22]. The frequency of ATRX mutations and their correlation to poor prognoses have spurred further exploration of the role of ATRX in PanNETs.

Interestingly, certain tumours with ATRX mutations exhibit heightened sensitivity to immune-mediated therapies (Fig 3). Specifically, ATRX loss can activate IRF transcription factors, leading to increased recruitment of CD4+ and CD3+ T-cells; however, concurrent IDH mutations may obscure this immune sensitization [18]. ATRX disruption has also been observed to enhance the cytotoxic effects of temozolomide (TMZ) treatment in gliomas by reducing ATRX-dependent inhibition of *FADD* transcription. FADD sequesters PARP from DNA, exposing the genome to TMZ and increasing effectiveness of treatment. Loss of ATRX expression is thus associated with increased susceptibility to radiation, chemotherapeutic agents, and immunotherapies, positioning ATRX as a potential therapeutic target [2,20,21].

Pituitary neuroendocrine tumours (PitNETs), traditionally known as pituitary adenomas, are common primary intracranial tumours of the adenohypophysis [45]. Like other neuroendocrine tumours, PitNETs are classified based on hormonal secretion. Overproduction and secretion of any adenohypophyseal hormones can cause debilitating effects associated with conditions such as acromegaly and Cushing’s disease. Although most clinically apparent PitNETs are benign, approximately 10% exhibit aggressive and invasive behaviour, leading to mass-effect symptoms from compression of adjacent intracranial structures [46,47]. Pituitary neuroendocrine carcinomas (PitNECs), formally pituitary carcinomas (PCs), account for only 0.1-0.5% of PitNET cases [46–50]. Metastases can occur in tumours that appear benign histologically while other tumours that have histological features of aggressiveness can remain isolated to the sellar region. Elevated mitotic activity and Ki-67 index scores are typically associated with aggressive pituitary tumours (APTs) and PCs [48,51]. These tumours often resist conventional surgical and radiotherapeutic interventions, with TMZ only showing moderate and usually transient benefits in less than half of patients [50,52,53]. Survival data for APTs and PCs indicate poor outcomes, with 3- and 5-year survival rates of 59% and 35%, respectively, and median survival times of 17.2 and 11.3 years for patients who are responsive to TMZ [48,54]. Alterations in genes regulating chromatin and genome stability – such as *USP8, USP48, BRAF, GNAS, and SF3B1* – are frequently associated with worse prognoses and shorter progression-free survival [55,56]. Despite various clinical and pathological markers, no definitive histopathological feature can reliably predict the transition from a benign to an aggressive or metastatic phenotype. Given the growing use of ATRX status as a prognostic marker in other neuroendocrine tumors, it is essential to explore whether ATRX may similarly predict PitNET aggressiveness and outcomes. The heightened susceptibility to mutagenesis and disrupted transcriptional regulation observed in ATRX-deficient gliomas, sarcomas, and other neuroendocrine tumours suggests that similar mechanisms could contribute to the aggressive and proliferative behaviours seen in rare PitNET subtypes.

Currently, published data on the incidence of ATRX expression and its clinical significance in neuroendocrine tumours, particularly PitNETs, is limited. The role of ATRX protein expression in the prognosis of PitNETs remains unknown. In this systematic review, we interpret the existing literature on ATRX expression in PitNETs to better describe its relationship with the clinicopathological characteristics and prognosis of PitNET patients.

## Methods

### Search strategy

This systematic review was conducted following the Preferred Reporting Items for Systematic Reviews and Meta-Analyses (PRISMA) guidelines. The aim was to identify all English published studies on pituitary tumours exhibiting a loss of ATRX expression. The strategy involved searching the PubMed, Embase, and Scopus databases to identify relevant studies published in English from January 1, 2014 to March 1, 2025, using MeSH terms combined with free search terms. A combination of the following keywords was used: ATRX, pituitary, pituitary adenoma, pituitary carcinoma, alpha thalassemia X-linked, ATRX, PitNET. No other restrictions were imposed, and no attempt was made to obtain unpublished results. All references and supplementary data from all selected articles were also considered. The review was not registered, nor was a review protocol prepared. All data collected and analyzed has been reported in the manuscript.

### Study selection

The selection of published material was conducted based on initial screening of titles or abstracts followed by screening of full-text reviews. Manuscripts were considered eligible if they met the following criteria: (i) the study was published in a peer-reviewed medical journal within the last ten years, (ii) the study was published in English, (iii) the study described human case studies involving pituitary tumours/PitNETs and ATRX, and (iv) the study observed a loss of ATRX expression or mutation in the ATRX gene (Fig 4). Studies were omitted from review based on the following exclusion criteria: (i) studies with publication types other than original research articles, such as review articles, editorials, letters, commentaries, and conference abstracts. Duplicate files were removed following export of database search results to Covidence. Risk of bias assessments were evaluated using the Joanna Briggs Institute (JBI) Critical Appraisal Checklist for Studies Reporting Prevalence Data and the JBI Critical Appraisal Checklist for Case Reports [57]. Quality assessment was not performed, and therefore no papers were excluded due to poor quality.

**Fig 4.**
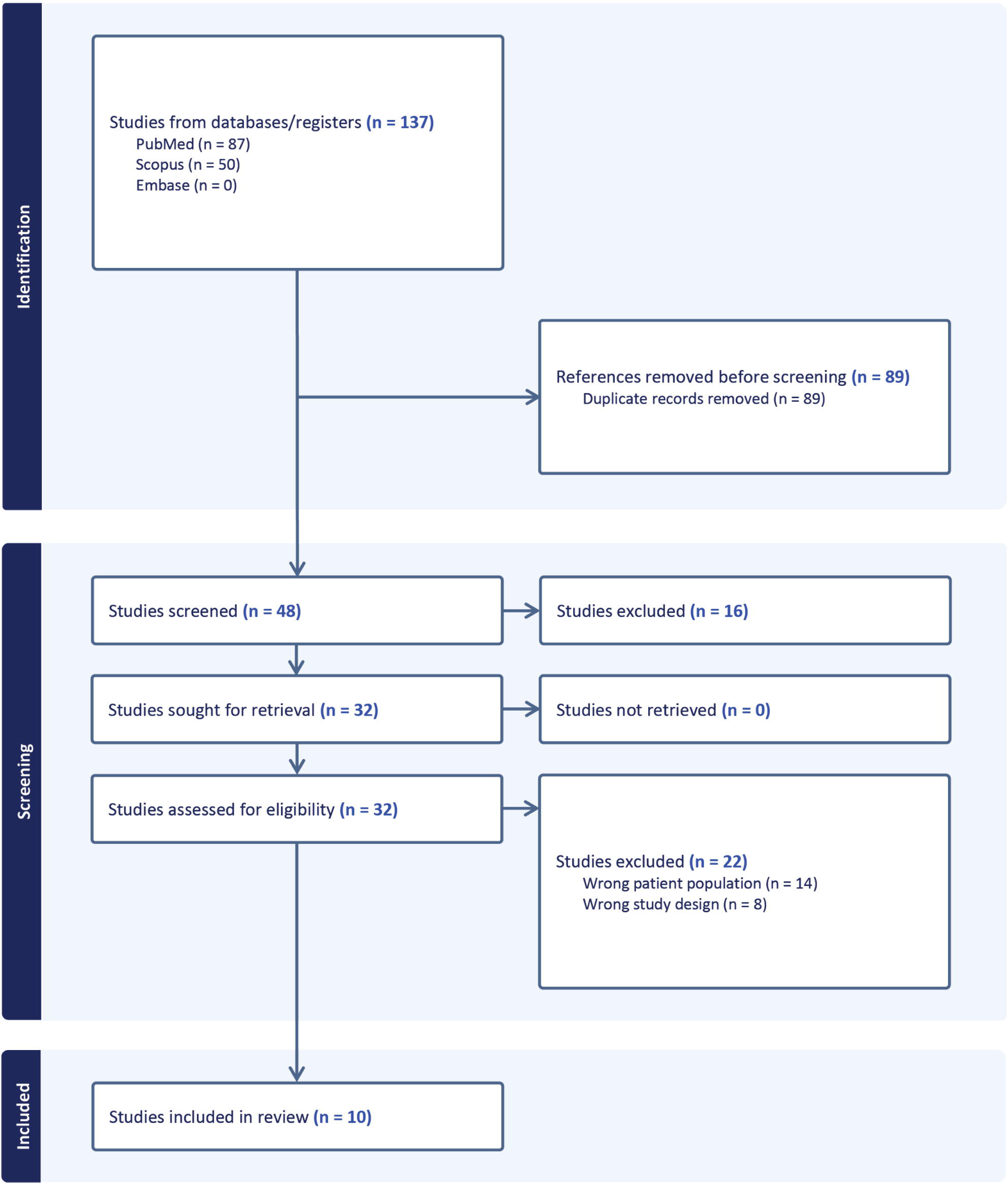
Identification and selection of studies via databases.

### Data extraction and quality assessment

Data was extracted from selected articles and included the following characteristics:

- Study: year of publication, journal of publication, country, type of study
- Sample Data: sample size, tumour type studied, mean age of participants at the time of tumour resection, sex, evaluation strategies,
- Study Results and Analysis: tumour classification, ALT status, Ki-67 index, identification of other mutations within tumour sample

The analyzed data, including methods and results of each full-length publication, were reviewed to satisfy the selection criteria and assess the quality of the text. No studies were excluded due to poor quality of methods or unsatisfactory results. Results were independently extracted by one author (EW) and assessed by another independent author (FR) to ensure accuracy and inclusion of complete data. Any discrepancies were resolved by discussion.

### Synthesis of data

All relevant patient demographics and tumour data were collected and tabulated in Excel®. Tumours that were referred to as ACTH-secreting tumours, ACTH-omas, or exhibited T-PIT expression were tabulated as corticotrophs. Null cell pituitary tumours were recorded as nonfunctional pituitary adenomas. Similarly, gonadotrophs included tumours categorized by LH/FSH secretion or SF1 expression. One study recorded gonadotrophs as nonfunctional pituitary adenomas and did not specify the number of adenomas recorded [58]. Pit-1 lineage tumours were further categorized into GH-secreting somatotroph and PRL-secreting lactotrophs unless not stated. One study published following the change in pituitary tumour classification also included acidophilic stem cell tumour classification, which was categorized as a lactotroph based on hormone produced [59]. Thyrotrophs included TSH-secreting adenomas and thyrotrophic adenomas. Any tumours that did not fall under these categories were tabulated under “Other Tumour Type,” including all plurihormonal tumours, pituitary carcinomas, and pituitary rhabdomyosarcoma. Tumours labelled as metastatic PitNETs were categorized as pituitary carcinomas for analysis. Tumours exhibiting ATRX within the included categories were indicated within square brackets. Analytical techniques and evaluation strategies were indicated in Table 2 while data that was unspecified or not measured was recorded in Table 3. The statistical analysis included Fisher’s exact test for associations between tumour recurrence, and other tumour characteristics and ATRX loss. No other statistical or sensitivity analyses were conducted.

**Table 2.**
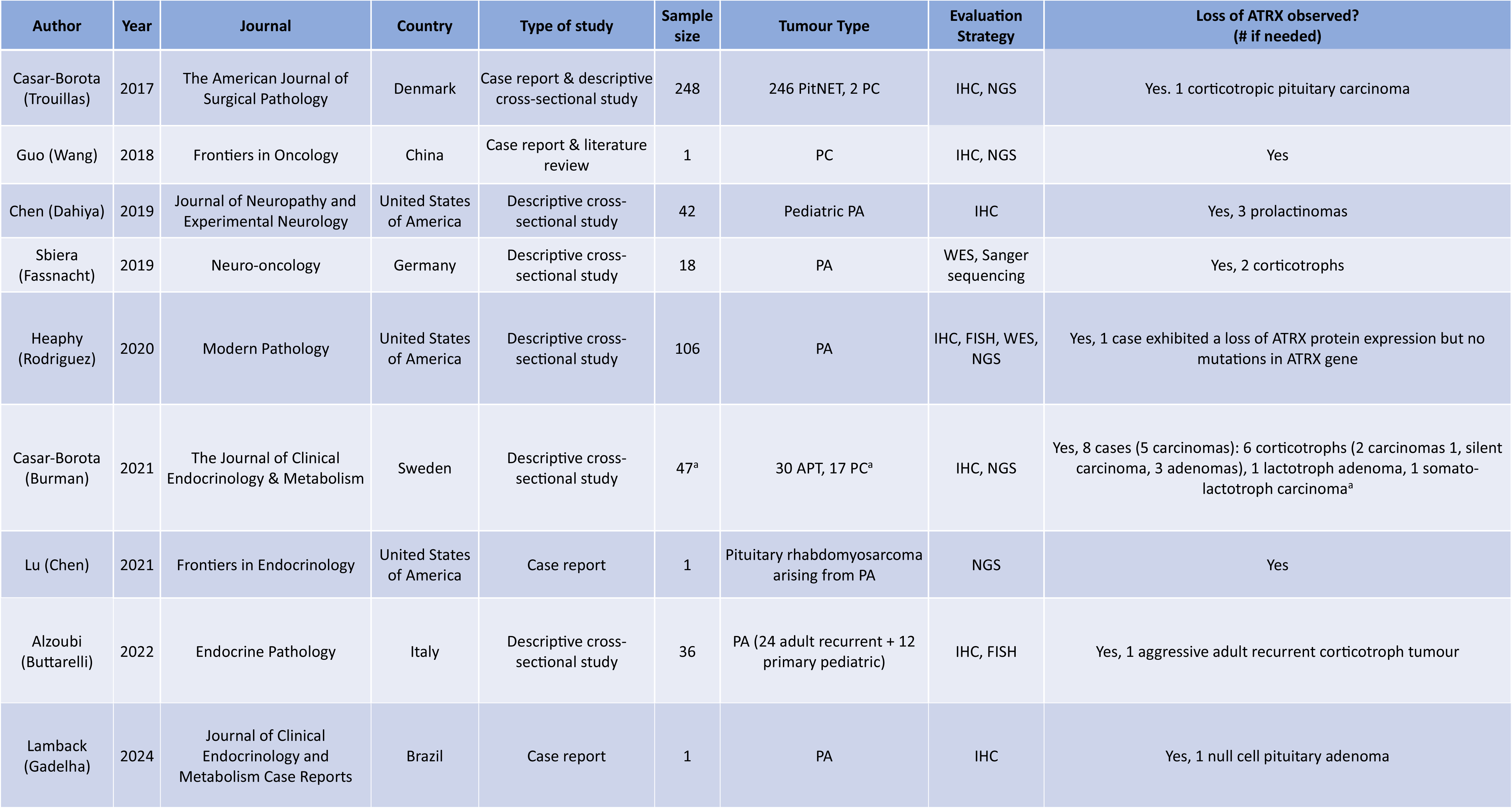
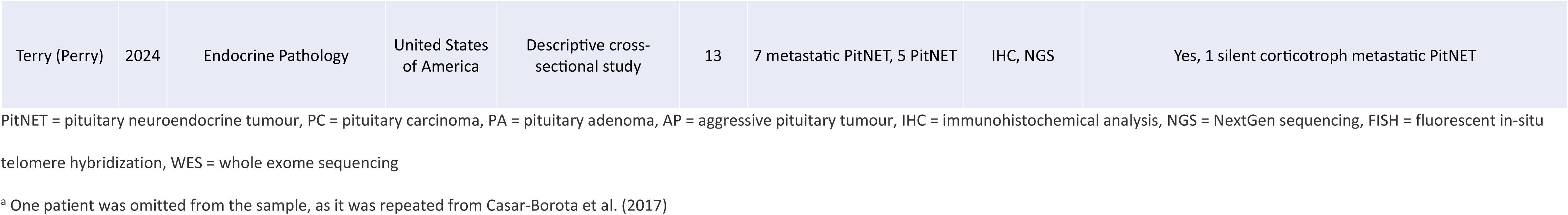
Characteristics of included publications examining the prevalence of ATRX loss in human pituitary neuroendocrine tumours.

**Table 3.**
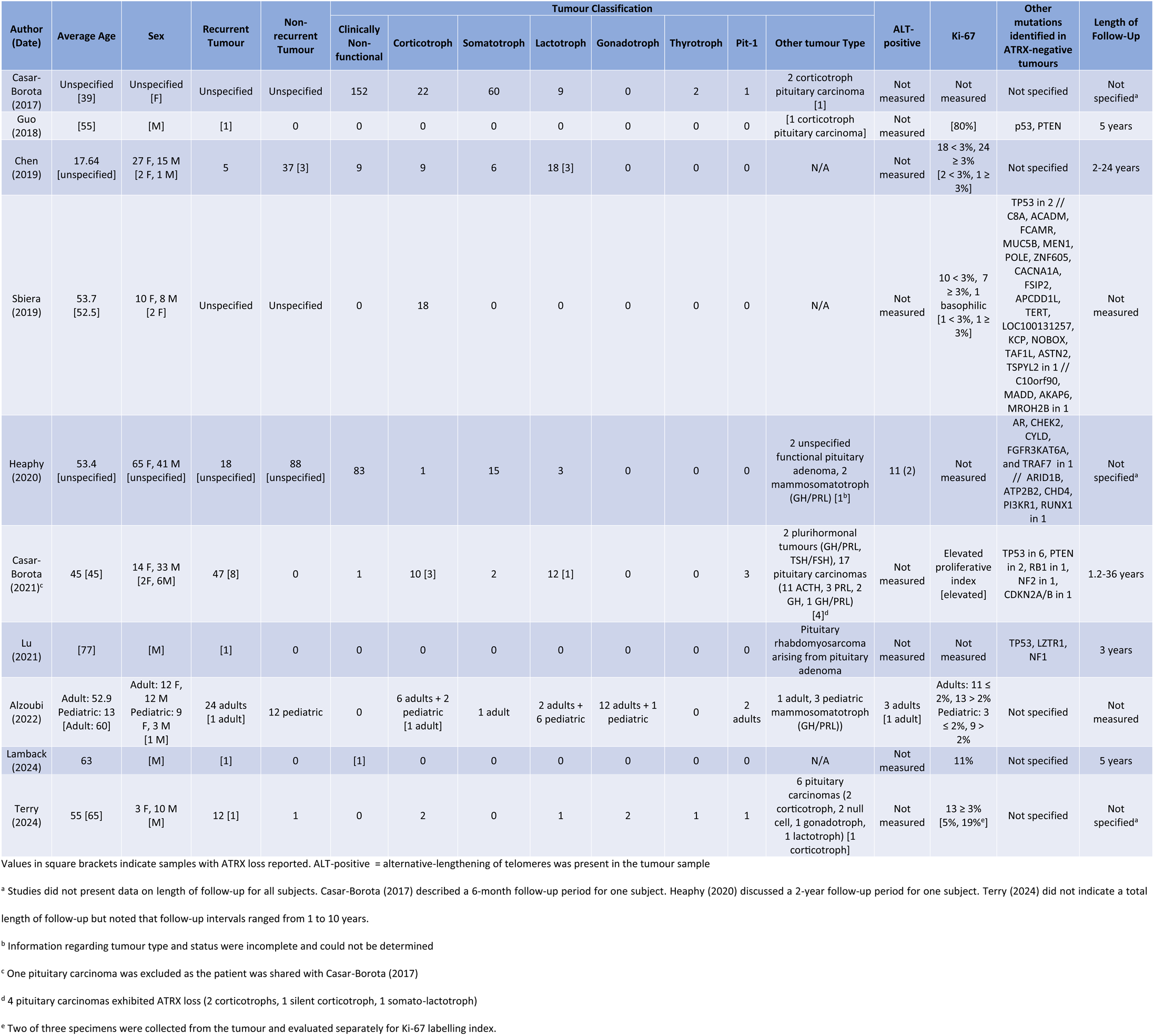
Demographic and clinical patient data examined in the included publications.

### Potential for bias and risk of bias assessment

Preliminary screening was accomplished by one of the authors (EW) and then reassessed by a second author (FR) according to the defined criteria to ensure that the same data items were collected. The information was extracted from the full text independently by each author. These efforts were directed toward conducting the analysis with minimal bias. Risk of bias and quality assessments were determined using the Joanna Briggs Institute (JBI) Critical Appraisal Checklist for Studies Reporting Prevalence Data (see Table 4) and the JBI Critical Appraisal Checklist For Case Reports (see Table 5) [57].

**Table 4:** Quality assessment using the Joanna Briggs Institute (JBI) Critical Appraisal Checklist for Prevalence Studies.

|  | Casar-Borota (2017) | Chen (2019) | Sbiera (2019) | Heaphy (2020) | Casar-Borota (2021) | Alzoubi (2022) | Terry (2024) |
| --- | --- | --- | --- | --- | --- | --- | --- |
| Was the sample frame appropriate to address the target population? | Yes | Yes | Yes | Yes | Yes | Yes | Yes |
| Were study participants sampled in an appropriate way? | No | Yes | No | No | Yes | Yes | Yes |
| Was the sample size adequate? | Unclear | Unclear | Unclear | Unclear | Unclear | Unclear | Unclear |
| Were the study subjects and the setting described in detail? | No | Yes | Yes | No | Yes | Yes | Yes |
| Was the data analysis conducted with sufficient coverage of the identified sample? | Unclear | Yes | Yes | No | Yes | Yes | Yes |
| Were valid methods used for the identification of the condition? | Yes | Yes | Yes | Yes | Yes | Yes | Yes |
| Was the condition measured in a standard, reliable way? | Yes | Yes | Yes | Yes | Yes | Yes | Yes |
| Was there appropriate statistical analysis | No | Yes | Yes | Yes | No | Yes | Yes |
| Was the response rate adequate, and if not, was the low response rate managed appropriately? | N/A | N/A | N/A | N/A | N/A | N/A | N/A |
| Final Scores | 3/8 | 7/8 | 6/8 | 4/8 | 6/8 | 7/8 | 7/8 |

N/A = Not applicable; the checklist item was not applicable to the study design
Studies with final scores $\geq 6$ are reported as “great” quality. Studies with final scores $\leq 4$ are reported as “poor” quality. Studies with final scores = 5 are reported as “moderate” quality.

**Table 5:** The Joanna Briggs Institute (JBI) Critical Appraisal Checklist for Risk of Bias for Case Reports that were identified.

|  | Casar-Borota<br>(2017) | Guo<br>(2018) | Lu<br>(2021) | Lamback<br>(2024) |
| --- | --- | --- | --- | --- |
| Were patient’s demographic characteristics clearly described? | Yes | Yes | Yes | Yes |
| Was the patient’s history clearly described and presented as a timeline? | Yes | Yes | Yes | Yes |
| Was the current clinical condition of the patient on presentation clearly described? | Yes | Yes | Yes | Yes |
| Were diagnostic tests or assessment methods and the results clearly described? | Yes | Yes | Yes | Yes |
| Was the intervention(s) or treatment procedure(s) clearly described? | Yes | Yes | Yes | Yes |
| Was the post-intervention clinical condition clearly described? | Yes | Yes | No | Yes |
| Were adverse events (harms) or unanticipated events identified and described? | Unclear | Yes | Unclear | Yes |
| Does the case report provide takeaway lessons? | Yes | Yes | Yes | Yes |
| Final Score | 7/8 | 8/8 | 6/8 | 8/8 |

Studies with final scores $\geq 6$ are reported as “good” quality. Studies with final scores $\leq 4$ are reported as “poor” quality. Studies with final scores = 5 are reported as “moderate” quality.

## Results

### Description of studies

A total of 48 studies were identified from database search after duplicates were removed. Following screening of titles and abstracts, 16 irrelevant studies were excluded. All 32 of the remaining studies were retrieved and assessed for inclusion. Of these, 22 publications failed to meet our inclusion criteria and were subsequently excluded. Fourteen studies were excluded due to wrong patient population, of which eleven studies did not examine samples including PitNETs for ATRX expression and mutations, while three studies were excluded due to ATRX mutations occurring outside of the pituitary tumour. Additionally, eight studies were excluded due incorrect study design, as the studies lacked inclusion of novel cases. Consequently, a total of ten publications were included based on the predetermined criteria [58–67].

Quality assessments were conducted for the nine studies deemed eligible for inclusion in our review study. Given that the selected descriptive cross-sectional studies were retrospective, analyzing tissue samples and existing medical records, subject response rate was not applicable and thus excluded from the risk of bias assessment. Accordingly, both case reports and descriptive studies were evaluated using an 8-point scale across various checklist parameters [57]. Of the ten human studies that analyzed the prevalence of ATRX loss in PitNETs, three studies presented case reports of a novel case that exhibited ATRX loss, while six were descriptive cross-sectional studies. One study presented both a novel case report and a descriptive cross-sectional study and was evaluated under both quality assessments [58]. Of the seven descriptive studies included in the review, 5/7 (71%) studies received “great” scores (≥6), while 2/7 (29%) studies were categorized as “poor” quality (<4) [58,63]. All four case reports received “great” scores. Nonetheless, because case reports generally focus on single patients and thus have limited generalizability and application, they still represent the weakest form of patient-based evidence (see Tables 4 and 5).

Table 2 summarizes the characteristics of the studies examined, including demographic data, sample size, and evaluation strategies. All studies examined were published since 2017. A study was published in each of Canada, China, Denmark, Germany, Italy, Sweden, and Brazil, and four studies published in the USA. Nine studies conducted immunohistochemical analysis to determine ATRX protein presence, while four studies conducted NextGen sequencing to determine the presence and location of ATRX gene mutations. One study did not conduct either immunohistochemical analysis or NextGen sequencing. Instead, the study examined selected tumours using whole-exome sequencing and Sanger sequencing [62]. One study also conducted NextGen sequencing to identify mutations in target genes, but did not report any sequencing results for the tumour lacking ATRX expression [59].

As 7/10 (70%) of the studies were conducted on data collected prior to the change in pituitary tumour classification, tumours in these studies were referred to as pituitary adenomas. Five studies examined adult pituitary tumour patients [59,60,62,65,67], while one study examined pediatric pituitary adenoma patients [61]. Three studies included both adult and pediatric pituitary adenoma patients [63,64,66]. One study did not disclose the age range of the patients included in their study [58]. In total, 26 pituitary carcinoma patients were also included from four studies in addition to their pituitary adenoma patients [58–60,64]. Additionally, one study presented a novel case study regarding a pituitary rhabdomyosarcoma arising from a pituitary adenoma that was identified to have a mutation in ATRX [65].

Patient data from the studies were summarized in Table 3, including basic patient information and tumour types. A total of 513 tumours were examined across the ten studies included. 246 tumours were clinically non-functional adenomas, 84 somatotroph adenomas, 70 corticotroph adenomas, 51 lactotroph adenomas, 15 gonadotroph adenomas, 3 thyrotroph adenomas, and 7 pituitary adenomas present with Pit-1. Several plurihormonal tumours were also included and were described in Table 3. The studies also reported 27 pituitary carcinomas, including 16 classified as corticotroph, 4 as lactotroph, 2 as somatotroph, 2 as null cell, 1 as gonadotroph, and 1 as plurihormonal. It was noted that in 3 of the studies reviewed, data involving age (average), sex, and tumour recurrence was incomplete [58,62,63]. Of the reported ages, the weighted average age of patients was 52. Among the publications that included sex distribution data, there were 148 female patients and 118 male patients. Additionally, 109 tumors were identified as recurrent, while 139 were classified as non-recurrent. However, data on follow-up duration were incomplete, with only 5 out of 9 studies providing information on length of follow-up for all subjects. Among these studies, reported follow-up periods ranged from 1.2 to 36 years [60,61,64,65,67].

Histopathological data regarding all tumours exhibiting ATRX loss included in this review were summarized in Table 6. Only 20/513 (3.9%) tumours exhibited ATRX loss, of which 12/20 (60%) tumours were classified as corticotrophs while 4/20 (20%) were classified as lactotrophs. One study reported a loss of ATRX protein expression in one tumour and partial loss in four other tumours, but did not describe the patient or tumour type of the tumours that exhibited loss of ATRX protein expression [63]. Furthermore, the researchers sequenced only the tumour sample that exhibited full loss of ATRX expression and did not reveal any ATRX genetic abnormalities. They also did not evaluate further the four cases that showed partial loss, and no information was provided regarding the extent of the loss, so they were not classified as ATRX-negative tumors. 17/20 (85%) of the tumours lacking ATRX expression were collected from adult patients, with the remaining three reported as pediatric cases [61].

**Table 6.**
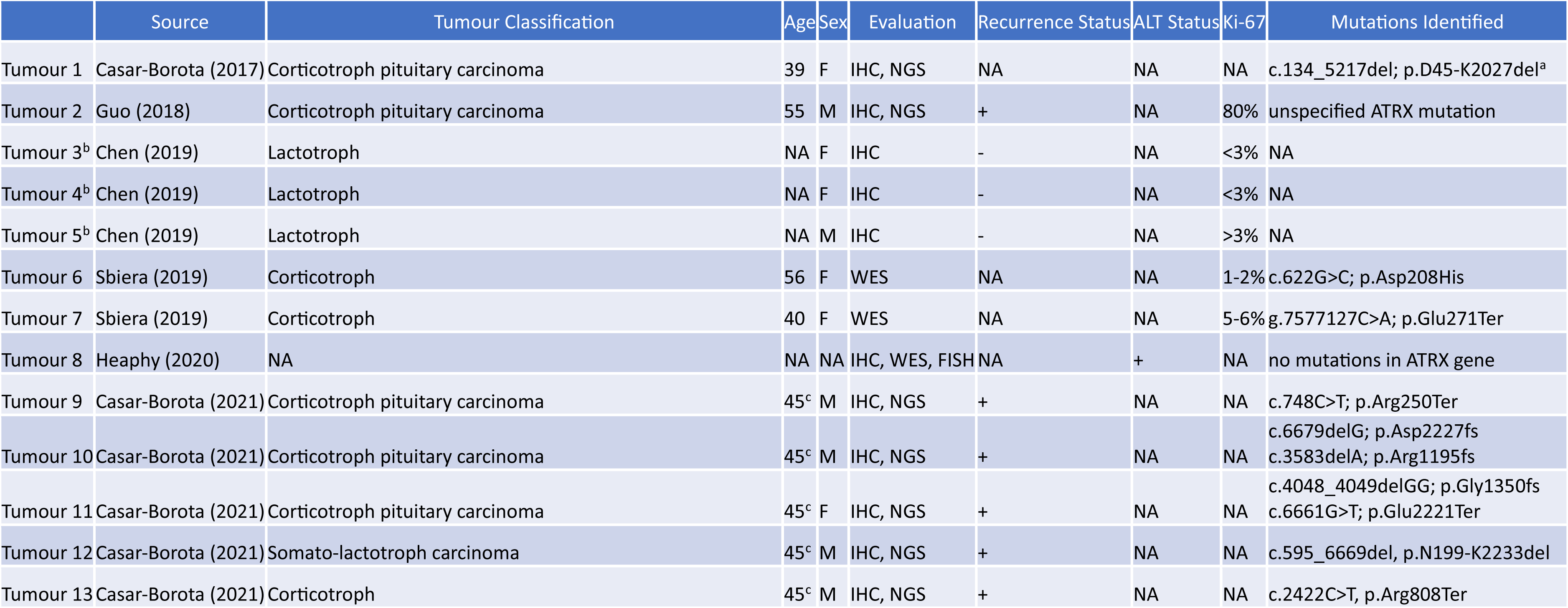
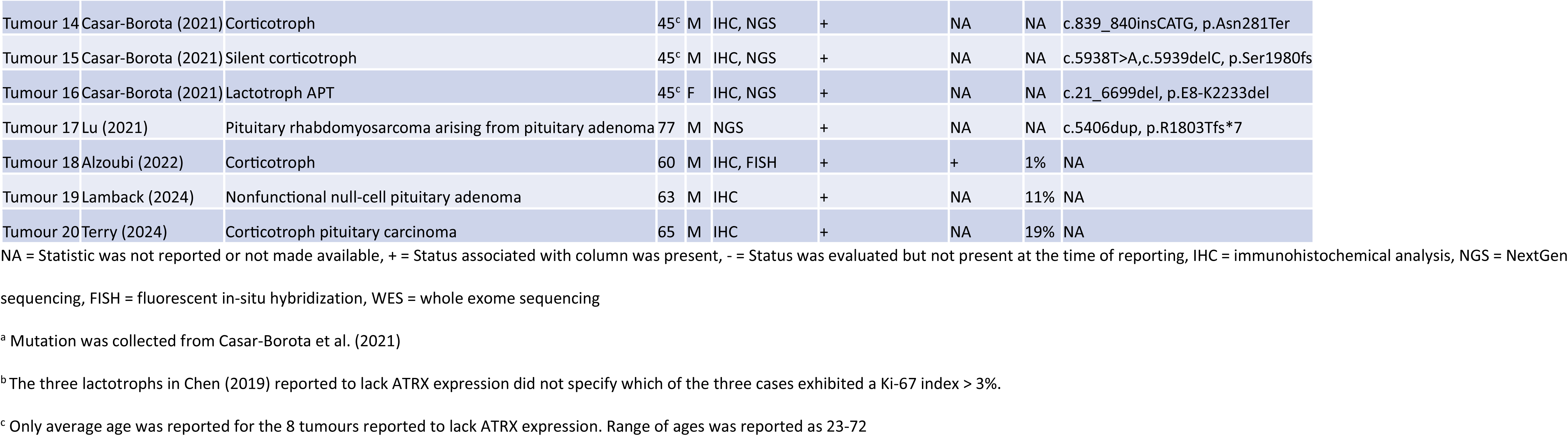
Demographic and histopathological characteristics of tumours exhibiting ATRX.

Two studies [63,65] reported the number of patients exhibiting the ALT phenotype, of which a total of 14 cases were identified and only 2/14 (14%) tumours exhibited ATRX loss. 5/9 studies reported the presence of other mutations within ATRX-negative tumours, of which *TP53* was the most frequently reported mutation (10/18). One study investigated the presence of 20 common glioma mutations, including IDH1, IDH2, and ATRX, within the analyzed pituitary tumours, of which 1/248 tumours exhibited a loss of ATRX due to a large deletion within the gene [58]. It was not specified, however, what the other 17 mutations the tumour was screened for or if any other mutations were observed. Ki-67 index was also reported by six studies [59–62,66,67] with different parameters, in which two studies reported proliferation indices greater than 3% as elevated [61,62], whereas another study reported indices greater than 2% as elevated [66]. One study reported an increase in their Ki-67 index by 80% from a prior examination but did not disclose what the absolute values were [60]. One study also reported indices greater than 10% as a marker of high proliferation but did not specify what they determined to be the threshold for an elevated index [67]. Various Ki-67 index thresholds for prognostic significance have been reported in the literature, generally ranging between 1% and 10%; hence, all thresholds utilized by the reviewed studies fall within the recognized prognostic range [68]. One study investigated aggressive pituitary adenomas and pituitary carcinomas, which were both characterized by increased proliferation [64].

Of the 26 pituitary carcinomas included in the review, 7/26 (27%) exhibited a loss of ATRX expression. 6/7 (86%) carcinomas with ATRX loss were corticotroph in nature while 1/7 (14%) presented as a somato-lactotroph. The remaining 19 pituitary carcinomas consisted of 10/19 (53%) reported to be corticotroph in nature, 3/19 (16%) as lactotroph in nature, and 2/19 (11%) as somatotroph in nature. The average age of pituitary carcinoma patients was between 16 and 73, with an average age of 46.1. Unfortunately, average age of carcinomas exhibiting ATRX loss cannot be calculated due to incomplete reporting by one study [64]. All carcinomas exhibited aggressive and transformative properties concomitant with their diagnosis, including elevated proliferative growth, need for repeated surgical procedures, resistance to drug therapies, and development of metastases. None of the studies that included pituitary carcinomas examined the presence of the ALT phenotype. Only two of the studies that presented pituitary carcinoma patients reported Ki-67 indices [59,60]. One study collected Ki-67 indices for two samples of the same tumour, recording 5% and 19% scores [59]. The second study reported an increase by 80% from a previous examination but did not provide information regarding the absolute value itself [60].

In the analysis of the relationship between ATRX loss and tumour recurrence [Table 7A], tumour hormone secretion [Table 7B], and patient sex [Table 7C], one study was excluded due to incomplete data regarding tumour recurrence, tumour type, and sex of patients with ATRX loss [63]. Two other studies were excluded from tumour recurrence and sex analysis due to incomplete reporting on the two parameters [58,62]. We did not find any significant relationship between loss of ATRX expression and tumour recurrence status [Table 7A]. Sex was also not a significant factor in ATRX expression [Table 7C]. However, tumour type was associated with ATRX expression status. As described in Table 7B, the proportion of ACTH-secreting tumours (corticotroph adenomas and corticotrophic pituitary carcinomas) exhibiting loss of ATRX expression was significantly different from the proportion of other tumours that exhibited ATRX loss (p < 0.0005). No other significant associations were observed.

**Table 7.**
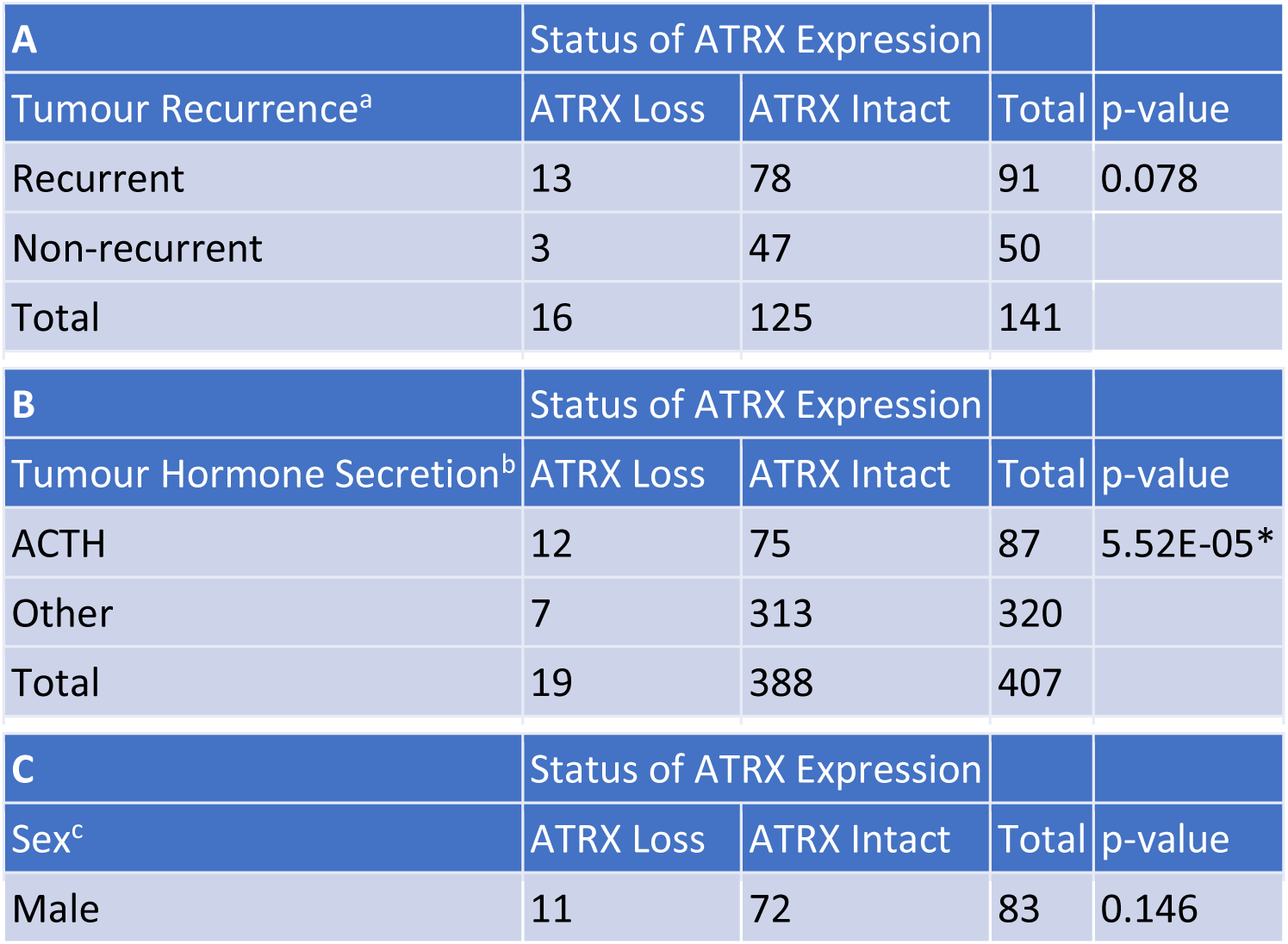
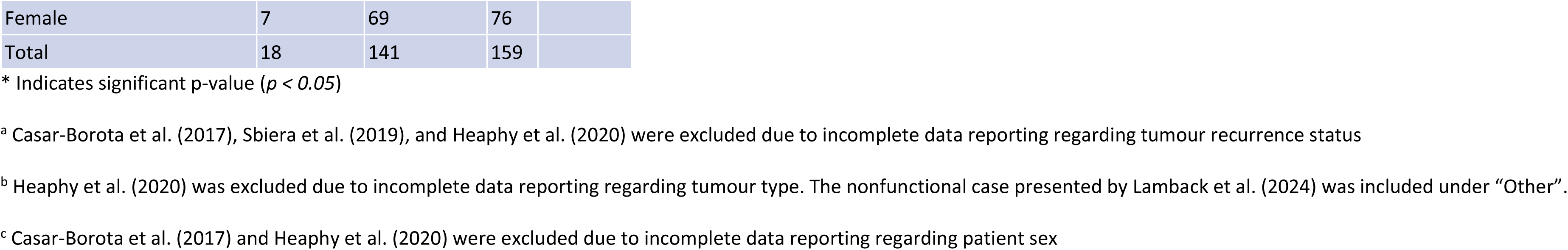
Fisher’s exact tests comparing ATRX expression status and tumour recurrence (A), tumour hormone secretion type (B), and sex of patient (C).

## Discussion

In the current study, we reviewed all available literature on ATRX expression in PitNETs. Our analysis revealed a loss of ATRX expression in approximately 3.9% of the 513 cases that were studied. Although ATRX is rarely mutated within PitNETs, patients with functioning PitNETs, especially corticotrophs, are over-represented (p < 0.0005).

We observed ATRX loss in 4 PitNETs exhibiting the ALT phenotype. However, not all ALT-positive tumours exhibited ATRX loss, suggesting that ATRX loss is not required for the ALT phenotype to progress within the PitNETs studied. ATRX-independent ALT phenotype progression has been observed in other tumour types, supporting this observation [69]. However, the limited examination of ALT among included studies limits further examination of this observation. While ATRX loss has been strongly associated with the ALT phenotype in tumour types such as gliomas and pancreatic tumours, we cannot determine a relationship between ATRX function and the progression of the ALT phenotype in PitNETs from currently published data.

Given ATRX’s critical role in sustaining methylation of repetitive DNA regions, a loss of ATRX expression may lead to errors in DNA replication and transcription during cell division. Coupled with an increased propensity for the development of the ALT phenotype, impaired ATRX function may promote the aberrant proliferative and transformative qualities in PitNETs observed in the analysis. Similar trends have also been identified in PanNETs, where the loss of ATRX/DAXX expression is associated with an increased propensity for proliferative development and poor survival [35]. All studies reviewed here consistently reported an increase in proliferative or transformative properties in tumours lacking ATRX expression. Four studies reported an elevated incidence of ATRX loss in recurrent or aggressive PitNETs, both of which presented with elevated proliferative traits [63,64,66,67]. However, given the multitude of factors, including initial degree of tumour resection and tumour location, the data we collected and analyzed from the publications demonstrated a non-significant trend with ATRX loss and recurrence in PitNETs.

Detailed mutation information was available for only 12 of 20 tumours lacking ATRX expression, with 11 cases exhibiting large mutations disrupting multiple domains of the ATRX protein, including frameshifts, early terminations, and extensive deletions. These broad mutations potentially disrupt ATRX-associated pathways, including PRC2- and PML-mediated chromatin regulation processes. Additionally, impaired ATRX-mediated sequestration of MRN complexes and compromised regulation of DNA damage responses may further increase susceptibility to mutations in other tumour suppressor genes or oncogenes, thereby driving aggressive and transformative tumour characteristics in the PitNETs discussed in this review.

Loss of ATRX expression was also noted in 27% of the 26 pituitary carcinomas presented across the studies included in this review. Pituitary carcinomas exhibit significant proliferative and transformative properties. The elevated prevalence of ATRX mutations among pituitary carcinomas supports the proposed association between ATRX loss and increased propensity for aggressive and proliferative development of tumours. It was also identified that 86% of the ATRX-negative carcinomas were corticotroph in nature, similar to the trends identified in the data from other ATRX-negative pituitary tumours collected in the review. However, as corticotrophic carcinomas were overrepresented across the samples, making up over 60% of all reported pituitary carcinomas, it is difficult to draw any conclusions from this observation. It is also important to note that in one excluded study, a pituitary carcinoma was found to exhibit ATRX mutations and expression loss in a liver metastasis but not in the original tumour, highlighting the potential for pituitary carcinomas to gain ATRX mutations [70].

It is worth noting that mutations in TP53, a well-established tumour suppressor gene, were identified in 50% of tumours exhibiting ATRX loss in the current review. As one of the most frequently mutated genes in cancer, TP53 mutants have a well-established association with proliferative development [71]. Notably, TP53 mutations have been proposed as markers to distinguish neuroendocrine carcinomas from other neuroendocrine neoplasms [72]. Concurrent mutations in TP53 and ATRX have been frequently observed in gliomas and sarcomas, with several studies suggesting the combination of these mutations as prognostic indicators associated with poorer survival outcomes [73,74]. The co-occurrence of TP53 and ATRX mutations presents a significant challenge in isolating and specifically assessing the impact of ATRX loss on the development and prognosis of PitNETs. Given the role of p53 in DNA repair and tumour suppression, mutations in TP53 may increase the propensity for ATRX mutations, thereby promoting proliferative development in pituitary tumours. Future studies may consider investigating ATRX expression in PitNETs and the interplay with TP53 mutations in prognosis of these patients.

All tumours exhibiting ATRX mutations and loss discussed in this review were identified using either immunohistochemical analysis (IHC), next-generation sequencing (NGS), or a combination of both. Although no studies have directly compared the effectiveness of NGS and IHC in identifying and assessing ATRX loss in samples, both methods are well-established and widely utilized in the field. The use of IHC for pathological analysis is a standard practice for endocrine and neurological tumour diagnoses as a relatively inexpensive means of detecting changes in protein expression. As ATRX mutations commonly result in truncated proteins, IHC is used to identify loss of ATRX protein expression common in several tumour types [75,76]. However, certain ATRX mutations have been found to result in false ATRX positivity during IHC due to positive staining for protein expression despite loss in protein function [77]. As IHC is limited by the antibodies available for detection, unusual mutations can limit the accuracy and effectiveness of such analyses for ATRX mutations. Furthermore, there is also a lack of standardization regarding determination of ATRX loss in tumours like gliomas, where specific thresholds for positive ATRX expression detected by IHC are not well enforced and vary between studies [78]. NGS allows for more comprehensive detection of tumour mutations and can detect multiple mutations across many genes in a single analysis. However, NGS is more costly and time intensive, making it less accessible [79]. NGS may also miss mutations outside of the gene that are impairing ATRX expression. Heaphy et al. identified a loss of ATRX expression in one case by IHC but did not identify any mutations from NGS, suggesting an external mechanism affecting ATRX expression [63]. A combination of IHC and NGS should be applied to provide comprehensive tumour examination. Three studies also used a combination of IHC, fluorescence in-situ hybridization (FISH), and whole exome sequencing (WES) for detection of ATRX mutations [62,63,66]. While FISH offers a cost-effective means of detecting large mutations, it is not sensitive enough to detect small intergenic changes that may affect ATRX expression [79]. WES applies similar practices to NGS, but instead examines a sample’s entire exome sequence, leading to far higher costs and time detection [77]. Thus, both FISH and WES are not as effective for frequent detection of ATRX mutations in PitNETs.

### Limitations

The aim of this study was to review all published research on ATRX mutation and expression in pituitary neuroendocrine tumours. To do so we must accept that we are limited to the type and quality of the published articles from other leading researchers within the field. With regards to our study, we found that the main limitation we identified was the heterogeneity in reporting of results, hampering the ability to perform analyses regarding outcome. For example, one study [63] did not report the tumour type or demographic data of the tumours exhibiting ATRX expression loss, while two other studies [58,62] did not report the average age, sex, and tumour recurrence status of patients in the study. Information regarding duration of follow-up following prognosis and treatment were also incomplete across the study reported in this review. Six studies reported varying follow-up periods between 6 months to 27 years, with many limited by patient loss and time between evaluation and publication [59–61,64,65,67]. Two studies also reported on the recurrent tumour characteristics and did not describe any evaluations of ATRX expression in the initial tumour [63,66]. Two studies did not present follow-up reports and data of the pituitary tumours [58,62]. The limited data presented regarding past visits and follow-up periods for patients included in each study hinders investigations on links to aggressive behaviour and recurrence. As a diagnostic marker of aggressive behaviour is recurrence, limited reporting and length of follow-up results may be omitting potential tumours exhibiting ATRX loss.

Measurement variability also limited investigations, as the included studies used different parameter thresholds for evaluation. The lack of a standard for declaring ATRX loss and the limited reporting on thresholds used during IHC affect the validity of IHC evaluations of ATRX loss. Furthermore, reporting on proliferative markers like Ki-67 were inconsistent, with varied standards for elevated values. As mentioned earlier, two studies reported elevated Ki-67 as values greater than 3% [61,62], while one study reported values greater than 2% as being elevated [66]. The inconsistent reporting of measurements limited any analyses and investigations. Evaluation of reported measures was also limited. While co-occuring mutations were observed and reported among tumours exhibiting ATRX loss, no further analyses were conducted to examine whether any other mutations affected tumour outcomes and measurements. Additionally, the limited research available regarding ATRX expression loss in pituitary tumours further restricted the statistical power of the review, as very few centres were involved in the collection of pituitary tumour samples used in the eight studies examined in the review. As most patients were collected from neurosurgical centres, patient data may be biased towards more severe and aggressive tumour cases.

### Future research

Our literature search revealed that all identified studies were published since 2017, highlighting this as an emerging field that requires further investigation. By comparing larger cohorts of PitNETs with ATRX loss to those with intact ATRX function, along with adequate long-term follow-up on outcomes, we can better understand the clinical significance of ATRX loss in PitNETs. Multivariate analyses of ATRX mutations against other observed mutations like TP53 will assist in isolating the effects of ATRX mutations on PitNET progression. Molecular analyses focused on ATRX-related pathways in ATRX-deficient PitNETs may also reveal deeper insights into the specific consequences of ATRX loss.

### Conclusions

Both ATRX loss and mutations are uncommon in PitNETs but when they do occur, they primarily affect functional tumours, particularly those of corticotroph nature. Furthermore, tumours lacking ATRX expression exhibit increased proliferative or transformative characteristics, including a higher incidence of ATRX loss in pituitary carcinomas. Although ATRX is associated with ALT phenotype repression, its loss was not found to exclusively dictate ALT phenotype occurrence, suggesting a more complex relationship between ATRX expression and ALT phenotype activation in PitNETs. This is an emerging area of study, and larger, prospective studies with long follow-up and consistent reporting are crucial to further investigate the implications of ATRX expression loss in PitNETs.

## Data Availability

All data for this study were obtained from publicly available sources cited in the manuscript. Extracted and summarized data supporting the findings of this study are included within the manuscript and Supporting Information files. No new data was generated.

## Data Availability

All relevant data are within the manuscript and its Supporting Information files.

## Acknowledgment

Authors are grateful to the Lloyd Carr-Harris Foundation for their continuous support.

## Notes

### Competing Interest Statement

The authors have declared no competing interest.

### Funding Statement

The author(s) received no specific funding for this work.

### Summary of Updates

Inclusion of three new figures and introduction. Updated systematic review results up to March 2025. Additional edits and revisions for clarity and depth.

